# Neurite orientation dispersion and density imaging in cocaine use disorder

**DOI:** 10.1101/2020.10.28.20221911

**Authors:** Jalil Rasgado-Toledo, Apurva Shah, Madhura Ingalhalikar, Eduardo A. Garza-Villarreal

## Abstract

Cocaine use disorder (CUD) is characterized by compulsive searching for cocaine that produces cognitive deficits, including lack of inhibition and decision-making. Several studies have shown that cocaine users exhibit brain volume and diffusion-based white-matter alterations in a wide variety of brain regions. However, the non-specificity of standard volumetric and diffusion-tensor methods to detect structural micropathology may lead to wrong conclusions. To better understand microstructural pathology in CUD, we analyzed 60 CUD participants (3 female) and 43 non-CUD controls (HC; 2 female) retrospectively from our cross-sectional Mexican SUD neuroimaging dataset (SUDMEX-CONN), using multi-shell diffusion-weighted imaging and the neurite orientation dispersion and density imaging (NODDI) analysis whose aims to more accurately model micro-structural pathology. We used Viso values of NODDI that employ a three-compartment model in white (WM) and gray-matter (GM). These values were correlated with clinical measures, including psychiatric severity status, impulsive behavior and pattern of cocaine and tobacco use in the CUD group. As hypothesized, we found higher whole-brain microstructural pathology in WM and GM in CUD patients than controls. ROI analysis revealed higher Viso-NODDI values in superior longitudinal fasciculus, cingulum, hippocampus cingulum, forceps minor and Uncinate fasciculus, as well as in frontal and parieto-temporal GM structures. Correlations between significant ROIs showed a dependency of impulsivity and years of cocaine consumption over Viso-NODDI. However, we did not find correlations with psychopathology measures. Overall, microstructural pathology seems to be present in CUD both in gray and white-matter, however their clinical relevance remains questionable.

## Introduction

Cocaine use disorder (CUD) is a worldwide substance use disorder (SUD) with serious consequences for the individual’s health and society, as it produces cognitive deficit, including lack of inhibition and compulsive search for cocaine, that may be secondary to brain pathology (Volkow et al., 2019) and, as of today, there are no proven successful treatments (Gorelick, 2015). The microstructural pathology in CUD commonly observed in postsynaptic receptors and dendritic spines, appears to be a consequence of neuroadaptation in neurons located in ventral tegmental area (VTA) and nucleus accumbens (NAc) (Siciliano et al., 08/2015). Using MRI in humans and rodents, researchers have observed macroscopic volume changes in regions of the frontostriatal and mesolimbic systems as well as cerebellum, though these changes are inconsistent between studies (Moreno-López et al., 10/2012; Narayana et al., 10/2010). While several studies show reduced volume, others show increased volume in regions such as basal ganglia and thalamus (Garza-Villarreal et al., 2017). This has also been observed with other morphometric measures such as cortical thickness, which has been shown to differ between studies as well (Moreno-López et al., 10/2012; Narayana et al., 10/2010). Elements contributing to this variability may be related to: 1) CUD pathology itself, 2) the unspecificity of volume or cortical thickness as macrostructural measures, and 3) the moment in time when volume was measured, as it also contributes to volume variability, among others. Of these elements, the inclusion of other whole-brain structural measures may uncover otherwise unknown underlying microstructural adaptations in vivo.

Diffusion weighted imaging (DWI) is an MRI modality that may help capture microstructural pathology and is a candidate for improving in vivo measurements. However, DWI in CUD has also shown variability of the regions affected, the magnitude, and the type of pathology (Ma et al., 10/2009; Romero et al., 1/2010). The reasons seem to be similar to other structural measures: 1) the unspecificity of typical DWI measurements such fractional anisotropy (FA), mean diffusivity (MD), and tractography, and 2) CUD itself with varying chronicity of cocaine use and abstinence periods (van Son et al., 2016). A recent meta-analysis of stimulant use disorders showed a lack of consistency between DWI studies, illustrating small to moderate effects in WM FA values with common analysis in a small group of cocaine use studies, with large heterogeneity and population differences between them (Beard et al., 2019). One suggested method that could capture microstructural pathology in vivo is neurite orientation dispersion and density imaging (NODDI) (H. Zhang et al., 2012).

Fractional volume of extracellular fluid (Viso) from NODDI is a novel extension of the diffusion tensor imaging (DTI) model method that measures free-water molecules in gray and white matter using multi-tensor information. Compared with standard DTI measures, NODDI analysis has been shown to be more sensitive to detect disease specific effects such as WM deterioration, neuroinflammation, dynamics and conditions of cell formation processes (Grussu et al., 2017). These metrics are associated with clinical symptoms in pathologies such as schizophrenia, Alzheimer’s disease, Multiple sclerosis, and Parkinson’s disease (Dumont et al., 2019; Grussu et al., 2017; Mitchell et al., 2019). Therefore, the measurement of free-water by NODDI seems to be a state-of-the-art sensible marker of brain pathology and may provide insight on CUD pathology. In this study, we wanted to determine CUD structural pathology in the human brain using in vivo MRI and the NODDI technique, as well as its correlation with clinical measures.

## Materials and Methods

We recruited 74 cocaine use disorder patients (CUD) and 64 non-CUD controls (HC) for a substance use disorder database (SUDMEX). HC were matched by age (± 2y), sex, and handedness. Education was matched as closely as possible. The recruitment criteria are shown in Supplementary Table 1 while the clinicals instruments used for are shown in Supplementary Table 2. For this study, we included 60 CUD (3F) / 43 HC (2F) with a median age of 31 (18 – 50) years old from the SUDMEX dataset. We eliminated 33 participants (HC = 21, CUD = 12): eliminated 7 due to other substance use dependence background (HC = 6, CUD = 1), 4 due to cocaine recreational use (CUD), 1 due to illiteracy (CUD), 1 due to excessive movement or artefacts related to image acquisition (CUD), 2 due to health clinical record (HC = 1, CUD = 1), 11 due to mental disorder record (HC = 10, CUD = 1), and 9 due to DWI non-acquisition (HC = 4, CUD = 5). The study was approved by the local ethics committee and carried out at the Instituto Nacional de Psiquiatría “Ramón de la Fuente Muñiz” in Mexico City, Mexico, according to the Declaration of Helsinki. All participants were invited through posters placed in several centers for SUD treatment. HCs were recruited from the Institute (i.e. administrative workers) and by fliers around the city. Participants were fully briefed and they provided verbal and written informed consent. Tobacco use in CUD is unavoidable, therefore we determined years of tobacco use and tobacco dependency in CUD and HC. Psychiatric comorbidities, lifetime medication and polysubstance use are reported in Supplementary Tables 3, 4 and 5 respectively. Participants were asked to abstain from any substance use, for at least 24 hours prior to the study and were urine-tested for the presence of the drugs before the clinical interview and MRI scan. The clinical and MRI sessions were done the same day as minimum and 4 days apart as maximum.

**Table 1.** Participant data.

|  | CUD (n=60) | HC (n=43) | Stats |
| --- | --- | --- | --- |
| Age | 31.23 (18 - 50) | 30.86 (18 - 48) | $t = -0.251, p = 0.801$ |
| Education | Middle School | High School | $\chi^2 = 15.71, p = 0.003$ |
| Handedness ** | (n = 60) | (n = 43) | $\chi^2 = 0.453, p = 0.797$ |
| *Right* | 53 | 36 |  |
| *Left* | 4 | 4 |  |
| Ambidextrous | 3 | 3 |  |
| Onset age of consumption | (n = 59), 20 (12 - 41) | na |  |
| Years consuming | (n = 59), 10 (1 - 28) | na |  |
| Average consumption per intake ** | (n = 57) | na |  |
| < 0.8 gm | 5 | na |  |
| 1.6 - 2.4 gm | 13 | na |  |
| 3.2 - 5.6 gm | 26 | na |  |
| 8 - 9 gm | 6 | na |  |
| 9 - 10 gm | 3 | na |  |
| > 10 gm | 3 | na |  |
| Monthly expense in cocaine (USD)* | (n = 56), \$127.5 (± \$205.59) | na | |
| Tobacco dependency ** | (n = 52), 34 | (n = 41), 18 | $\chi^2 = 19.22, p < 0.001$ |
| Years of tobacco use * | (n = 52), 13.71 (± 8.03) | (n = 41), 6.39 (± 8.145) | $T = -4.33, p < 0.001$ |

*Median (min – max) for all except: \* = mean (sd), \*\* = count. CUD = Cocaine Use Disorder, HC = Healthy controls, n = due to absence of data we show each sample size per variable, na = not applicable, gm = grams, Stats = statistics, t = t-value, $\chi^2$ = chi-squared.*

**Table 2.**
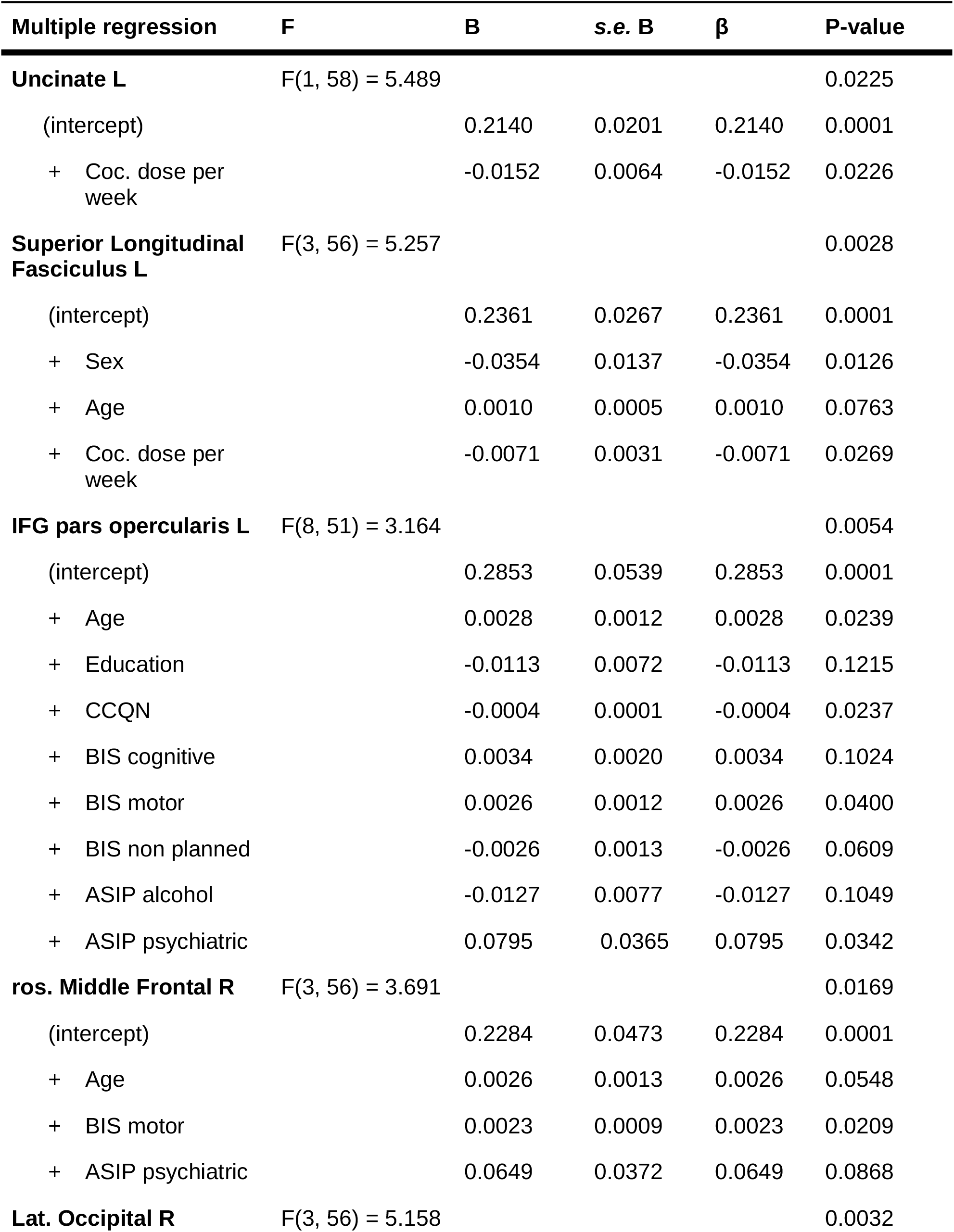
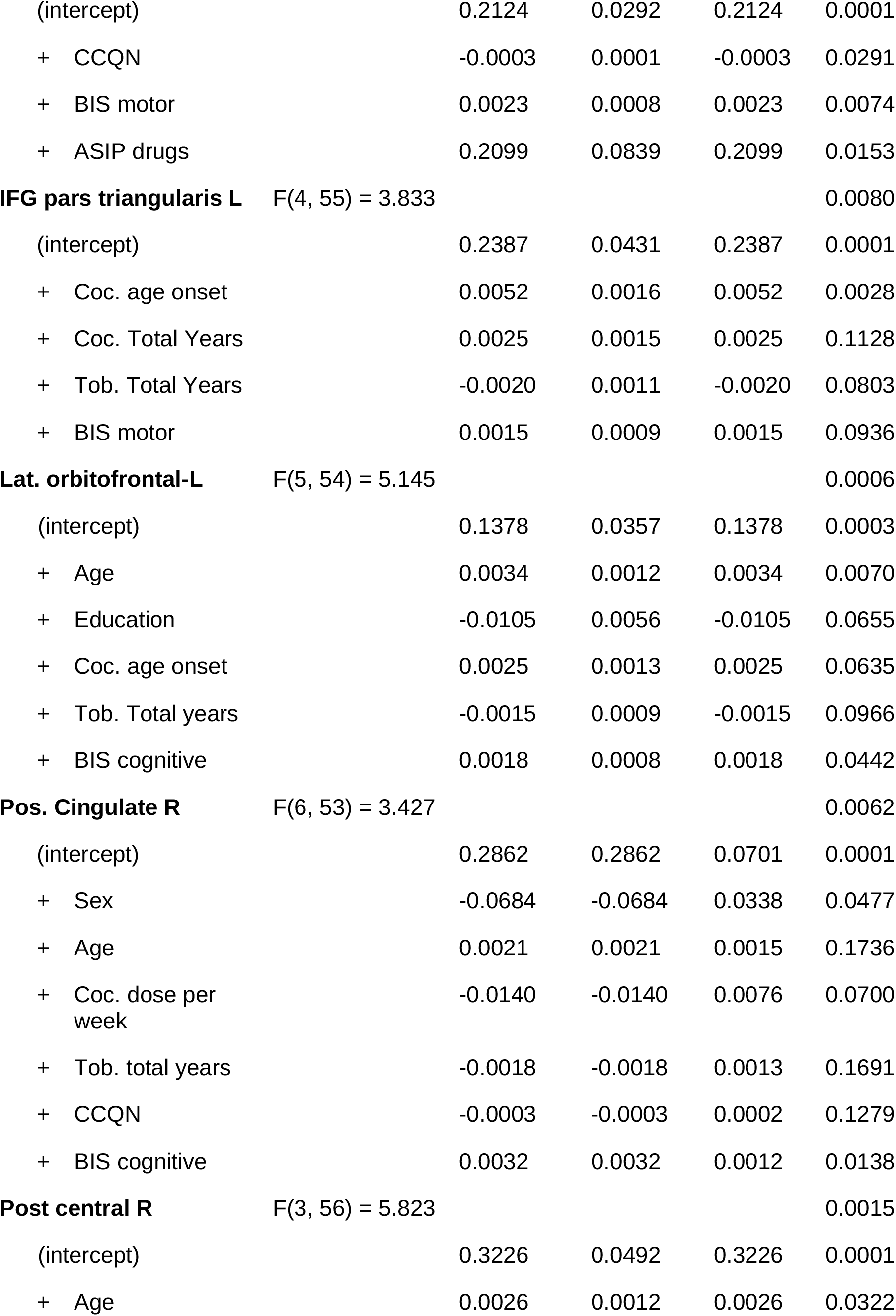
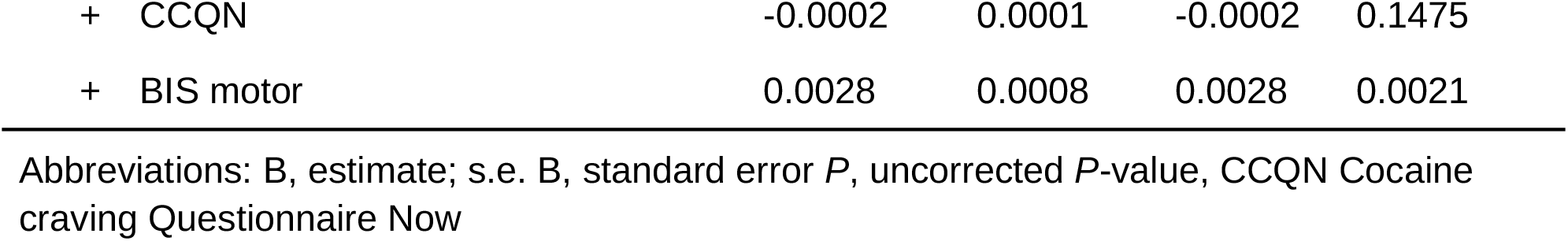
Multiple Regression between correlated brain regions and clinical measures.

### Clinical measures

Participants were asked to respond to an array of questionnaires in order to obtain CUD severity measures and tobacco use, administered by trained psychologists, psychology students and psychiatrists. For the CUD group, current craving at the time of MRI acquisition was evaluated using Cocaine Craving Questionnaire—Now (CCQ) which includes questions about desire, intent and planning to use cocaine, anticipation of positive outcome and of relief from withdrawal or dysphoria, and lack of control from using cocaine (Paliwal et al., 2008). Impulsivity was evaluated using the Barratt Impulsiveness Scale (BIS-11) that involves cognitive impulsivity which reflects attentional control, deliberate thinking and planning; non-planning impulsiveness which reflects a lack of forethought and motor impulsivity which reflects acting quickly and perseverance (Reise et al., 2013). Addiction Severity Index (ASI; (McLellan et al., 1985) were applied to assess substance use disorder diagnosis and severity of cocaine use.

### Magnetic resonance Imaging

Magnetic resonance images were acquired using a Phillips Ingenia 3T system (Philips Healthcare, Best, Netherlands & Boston, MA, USA) with a 32-channel dS Head coil. We acquired structural T1-weighted and DWI high angular resolution diffusion-weighted imaging (HARDI) images, as well as an opposite blip DWI sequence (AP). Specifically, T1-weighted images were acquired using a 3D FFE SENSE sequence, TR/TE = 7/3.5 ms, FOV = 240, matrix = 240 x 240 mm, 180 slices, gap = 0, plane = Sagittal, voxel = 1 x 1 x 1 mm (5 participants were acquired with a voxel size = .75 x .75 x 1 mm), scan time = 3.19 min. DWI HARDI used a SE sequence, TR/TE = 8600/127 ms, FOV = 224 mm, Matrix = 112 x 112, 50 slices, gap = 0, plane = axial, voxel = 2 x 2 x 2 mm, scan ∼20 min, directions 8 = b0, 36 = b-value 1,000 s/mm2 and 92 = b-value 3,000 s/mm2, total = 136 directions. As part of the SUDMEX dataset, resting-state fMRI and fast DKI sequences were also acquired and are not part of this particular study (Garza-Villarreal et al, 2017). The order of the sequences was: rsfMRI, T1w, HARDI, fDKI, and was maintained across participants. Total scan time was around 50 min.

## Data processing and analysis

### Image Processing

The preprocessing of DWI and T1 images included initial manual quality assessment and conversion from dicom to nifti format. The T1w images were then processed and parcellated into 86 regions of interest (ROIs) (68 cortical and 18 sub-cortical) of the Desikan atlas using Freesurfer 6.0 (Fischl, 2012). The pre-processing steps involved skull striping, bias correction, and tissue segmentation. Subsequently, surface-based non-linear registration to map the cortical sulci-gyri and volume-based registration for subcortical structures was performed (Fischl, 2012). These 86 ROIs were translated to the diffusion space by employing a symmetric diffeomorphic transformation model (SyN) using Advanced Normalization Tools (ANTs) (Avants et al., 2008). DWI volumes were preprocessed using FSL 6.0.11 (https://fsl.fmrib.ox.ac.uk). Initial steps included correction of eddy current artifacts and motion noise induced by eddy currents using “eddy_correct” which employed affine transformation to diffusion gradient images to the baseline b=0 image and distortion correction using “topup”. Based on the rotation parameters in the transformation, the gradients were rotated to match the transformed images. Subsequently, the brain extraction was performed using the brain extraction tool. To map WM regions we employed the JHU-WM atlas which contains 20 tracts defined in MNI space. Using ANTs registration, MNI space diffusion atlas (JHU atlas) was transformed into individual subject space to map all the ROIs into subject space.

We then used the neurite orientation dispersion and density imaging (NODDI) method that employs a three-compartment model to understand brain tissue microstructure in detail. NODDI produces three metrics that quantify: (1) the fractional volume of extracellular fluid (isotropic volume fraction [Viso]), (2) intracellular neurite dispersion (orientation dispersion index [ODI]) and (3) neurite density (volume fraction [Vic]), after eliminating effects of extracellular fluid. We employed the NODDI toolbox in MATLAB (H. Zhang et al., 2012) on preprocessed DWI images.

### Statistics

Statistical analysis was performed using SPSS software version 16.0 and R programming language v. 3.4.3. Participant demographics were compared between groups using analysis of variance (ANOVA). Distribution of sex was compared using a χ2 test. To examine the differences between NODDI metrics between CUD and HC groups, we performed a multivariate analysis of covariance (MANCOVA) test with age and sex as covariates. The test was carried out on mean Viso-NODDI values in parcellated gray matter (GM) cortical (68 ROIs) and subcortical (18 ROIs) regions as well as in white matter (WM) tracts and regions mapped from the JHU atlas. Significant group effects were adjusted for multiple comparisons using a 5% false discovery rate (FDR), followed by FDR-corrected pairwise comparisons. All reported *p*-values have been FDR corrected.

Only in CUD patients, the Viso-NODDI of every ROI were correlated with the addiction severity index (ASIP) scores, Barratt Impulsiveness Scale (BIS-11), Cocaine Craving Questionnaire (CCQ), consumption patterns and cocaine/tobacco pattern use. Partial Pearson’s r was computed, and the significance threshold of the correlation was maintained at an alpha = 0.05 with Education and Sex as covariables regressions. Also, global Pearson’s r was computed (Supplementary Figure 3). Finally, we corrected for multiple comparisons using false discovery rate (FDR) at 0.05 of p-values obtained. In order to determine the variables that most influence the Viso-NODDI values per significant ROI in the correlation, we used a stepwise regression with both forward and backward selection for searching the minimum Akaike Information Criterion (AIC) which provides a means for model selection, providing the loss of information for each model tested after removing the multicollinearity.

## Results

### Clinical characteristics

Clinical and demographic data characteristics summary of cocaine use disorder patients (CUD) and healthy controls (HC) are provided in Table 1. Chronological age was positively correlated with years of cocaine consumption (r = 0.63, p < 0.001), cocaine age onset (r = 0.45, p = 0.002) and years of tobacco consumption (r = 0.46, p = 0.002). CUD participants showed a significantly higher tobacco dependency (p < 0.001), years of tobacco use (p < 0.001) and a lower education (p = 0.003) when compared to controls. Also, years of cocaine consumption was positively correlated (r = 0.35, p = 0.022) with years of tobacco consumption (Supplementary Figure 1).

**Figure 1.**
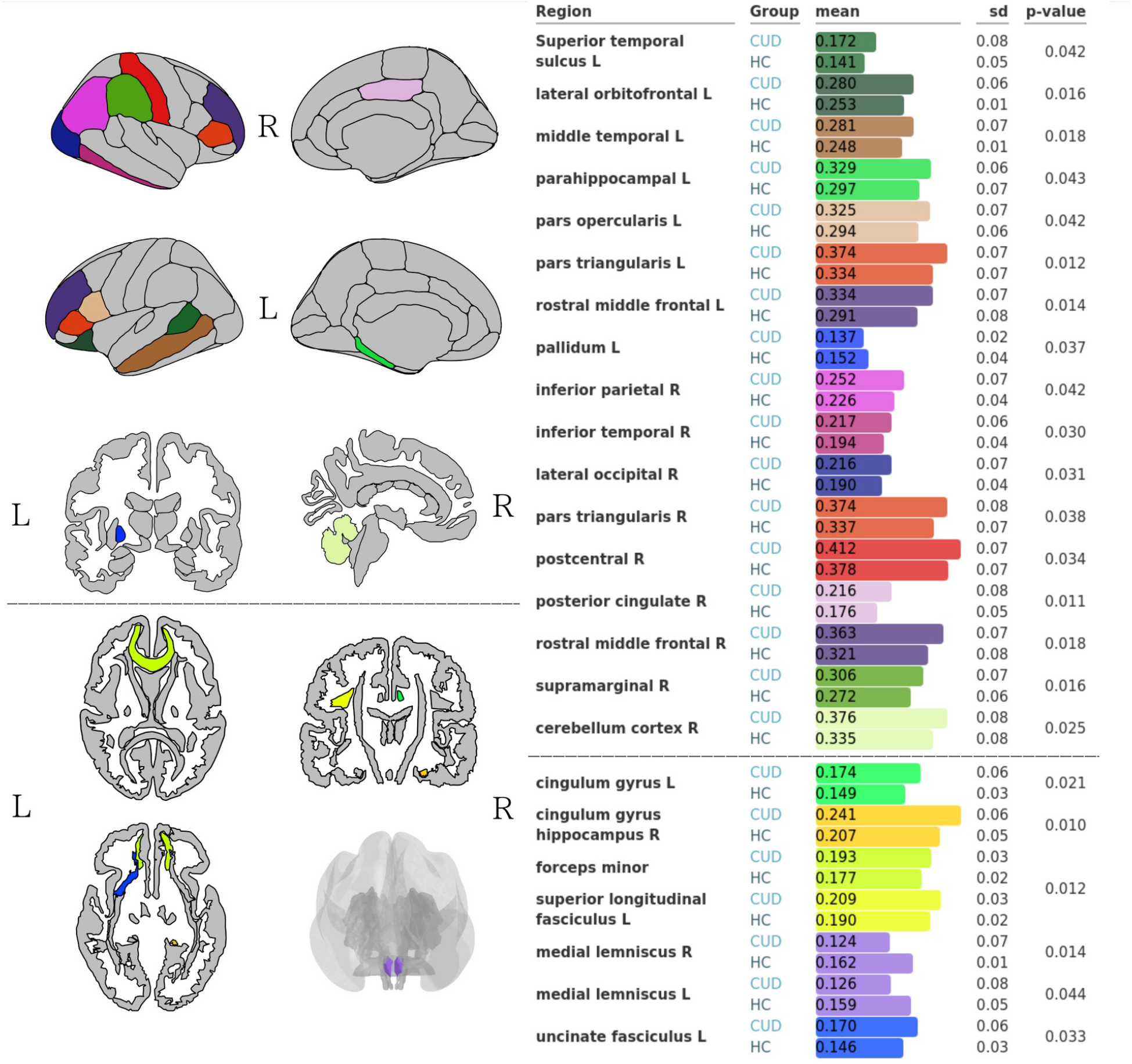
Viso-NODDI differences between groups in white and gray matter. Left: Gray matter regions (Desikan-Killiany atlas) and white matter regions (JHU WM tractography atlas) with Viso-NODDI statistical significant difference (FDR correction; p < 0.05). Right: Bar plots with mean Viso-NODDI values per significant region, standard deviation (sd) and adjusted p-values between cocaine use disorder patients (CUD) and healthy controls (HC). (Banks of) Superior temporal sulcus = bankssts L. Dashed lines indicate the separation between gray-matter and white-matter.

### NODDI analysis

We found higher mean Viso-NODDI values in WM and GM in the CUD group compared to the HC group in all ROIs (Supplementary Figure 2). In white-matter tracts (WM), Viso-NODDI values were significantly higher in CUD in some of the Regions of Interests (ROI) compared to HC in gray-matter and white-matter (Figure 1). For detailed results see Supplementary Tables 6 and 7.

**Figure 2.**
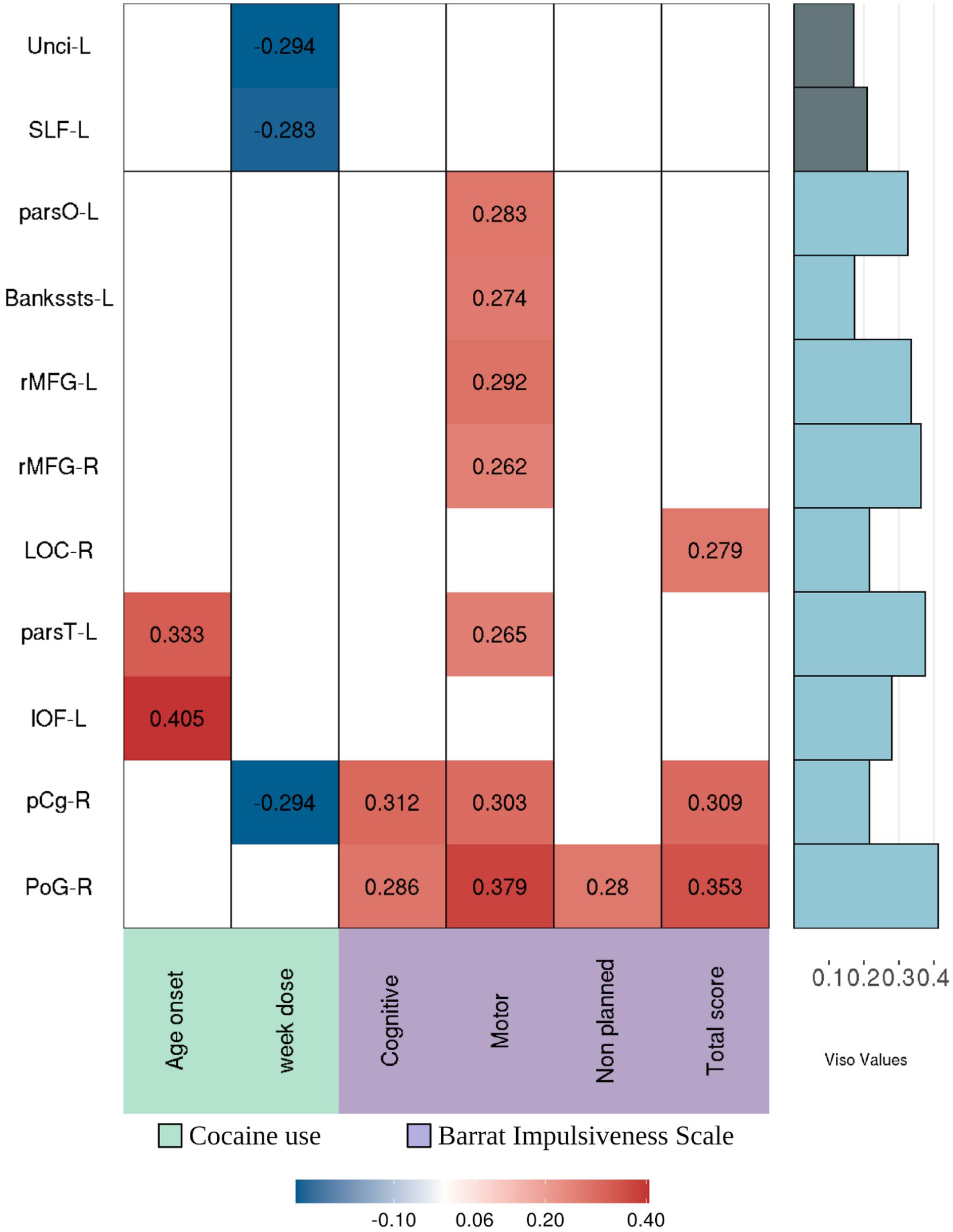
Partial correlation map between significant structures and clinical measures with education and sex as covariates. Partial correlation map between significant structures and clinical measures with education and sex as covariates. Blank space means no significance corrected after FDR (p<0.05) of significant values. Viso-NODDI values of each structure are plotted on the right side. Abbreviation: Unci-L = left Uncinate tract, SLF-L = Superior Longitudinal Fasciculus, parsO-L, Bankssts-L = left superior temporal sulcus, rMFG-L/R = left/right rostral Middle Frontal Gyrus, LOC-R = right lateral Occipital, parsT-L = Inferior Frontal Gyrus pars triangularis, lOF-L = left lateral Orbitofrontal Gyrus, pCg-R = right posterior Cingulate Gyrus, PoG-R = Right Postcentral Gyrus. Correlation details are present in Supplementary Table 8.

### Correlation of Viso-NODDI and clinical measures

CUD group correlation analysis between Viso-NODDI values and clinical measures revealed significant partial correlations after multiple comparisons correction (Figure 2). Positive correlations were found with impulsiveness index (BIS-11) in cognitive, motor, non planned and total score; cocaine age onset of consumption. On the other hand, cocaine dose per week was negatively correlated with WM structures and posterior cingulate (Supplementary Table 8).

The multiple regression results were an overall effect of sex, age, cocaine dosage per week and impulsivity on Viso-NODDI values in left Uncinate, left Superior Longitudinal Fasciculus, left Inferior Frontal gyrus pars triangularis and opercularis, left rostral Middle Frontal gyrus, right Lateral Occipital, right posterior Cingulate gyrus and right postcentral (Table 2).

## Discussion

In this study, we used NODDI imaging analysis to understand microstructural pathology in cocaine use disorder (CUD) patients. We found that: 1) CUD patients showed higher free-water (Viso-NODDI), both in bilateral gray matter structures and white matter tracts, except for right and left medial lemniscus where they show lower values than controls; and 2) several clinical substance use measures in the cocaine use disorder group correlated with viso-NODDI values. This study, to our knowledge, is the first to use NODDI imaging analysis in CUD patients where we found potentially affected brain regions.

General Viso-NODDI increments could be reflecting an increase in extracellular space free water that compromises cognitive performance like memory and executive functions, predisposing a general functional decline (Maillard et al., 2019). Although the underlying mechanism of these alterations remains unclear, studies have pointed out an important role of neuroinflammation, perturbed hemodynamic function, demyelination and axonal degeneration (Di Biase et al., 2020). These changes could be caused by genetic polymorphisms associated with different neurotransmitters that are implicated in drug consumption, previously described as a possible cause for WM changes (Azadeh et al., 2016). Cocaine consumption mediates microglial activation inducing inflammatory responses which may enhance the synaptic dopamine or glutamate concentrations, as reported in striatum of murine models (Burkovetskaya et al., 2020) and in white matter tracts in non-human primates brains, thus, contributing to the behavioral, cognitive and brain structural deficits (Smith et al., 2019).

### White matter pathology

The NODDI analysis showed global free-water-related pathology in white (WM) and gray matter (GM) in CUD patients. This type of pathology has been described in studies with other neuropsychiatric pathologies such as mild cognitive impairment, Alzheimer’s disease (Dumont et al., 2019) and Parkinson’s disease (Mitchell et al., 2019), suggesting Viso-NODDI’s sensitivity to structural pathology. Moreover, our region-of-interest analysis (ROI) revealed higher Viso-NODDI in 1) WM regions: left superior longitudinal fasciculus (SFL-L), right cingulum (Cg-R), right hippocampus cingulum (Cghi-R), forceps minor (FMi) and Uncinate fasciculus (Unci-L), and 2) GM regions: frontal and parieto-temporal cortices. Previous studies with single-tensor diffusion have shown that CUD is associated with higher or lower fractional anisotropy (FA) and radial diffusivity (RD) in widespread regions (Hampton et al., 2019). For example, studies have found reduced FA and increased RD in inferior frontal connections, orbitofrontal cortex (Lim et al., 2008), SFL, corticospinal tract, superiorcorona radiata, temporal and occipital lobes, cerebellum (Xu et al., 2010), thalamic radiation, internal capsule, and in corpus callosum; while higher FA has been also reported in anterior cingulate (Romero et al., 1/2010). Corpus callosum (CC) is the most consistently affected region found in the diffusion-weighted imaging (DWI) literature, and we found higher Viso-NODDI values in the FMi, which is part of the genu of the CC and connects the lateral and medial cortices between frontal lobes via the projections from anterior cingulate to anterior frontal regions (Ma et al., 10/2009). This suggests CC affectation may be related to interhemispheric frontal lobe pathology, which is highly implicated in executive function problems and cocaine craving (Ma et al., 10/2009).

We also found higher Viso-NODDI in PCC, Cg-R and Cghi-R. PCC is a core region hub of the default mode network (DMN), which is linked with emotional memory and social cognition (Downey et al., 2015), and has been implicated with worse recovery prediction from CUD treatment and with subjective craving in CUD (Wilcox et al., 2011). Moreover, the affectation of PCC suggests the DMN may be even more important to CUD pathology than previously thought, especially due to the involvement of another DMN core hub, the ventromedial prefrontal cortex, both connected by cingulum tracts (Greicius et al., 2009). (Reese et al., 2019) suggested that an imbalance between the DMN and the executive control network (ECN) through the modulation by salience network (SN) might be leading to a bias of internally oriented stimuli, such as withdrawal symptoms, where substance use severity is associated with reducing connectivity among these networks. This connectivity disequilibrium may provoke greater impulsivity and a compulsory seeking for substances, caused by the lack of response inhibition, relaxation in cognitive control and alteration in values of rewards, all regulated by structures into these networks and connected by genu of CC and Cg tracts (Dalley et al., 2011a). The higher Viso-NODDI found in cingulum (Cg-R and Cghi-R) could also be related to maladaptive behavior, affecting decision making, judgment, and planning (Bell et al., 2010). A similar role is played by superior longitudinal fasciculus which forms a link between parietal and temporal areas to prefrontal cortical regions (Makris et al., 2005), and in the same way that cingulum fibers, is implicated in emotional processing as well as memory and language, which are usually reported as disturbed in CUD subjects and prenatally exposed adolescents (Morie et al., 2017). Another relevant WM tract we found affected was the Uncinate fasciculus, which has been related to emotional empathy and stress due to the fronto-limbic connections between structures implicated on these processes, such the orbitofrontal cortex, anterior parahippocampal and amygdala (Oishi et al., 2015). Fractional anisotropy-based (FA) degeneration in this tract has been observed in brain injuries, behavioral disorders and psychopathy (J. Zhang et al., 2014), but also found in cocaine polysubstance users where it is described as part of the induced cocaine brain damage, reversed after an abstinence period (van Son et al., 2016). All these association fibers have in common the connection of structures that are responsible for emotion, social adaptation, superior behavioral control and decision making. An alteration of FA and RD values in these tracts has been found in some studies of cocaine use disorder (He et al., 2020). In this sense, Viso-NODDI may provide improved specificity of microstructural pathology in CUD that has been also observed in other studies with regular DWI.

### Gray matter pathology

We also found higher Viso-NODDI values in frontal, temporal and parietal gray matter, regions found to have reduced volume in cocaine users, involving overestimation of rewards, breakdown to inhibitory processes and impulsiveness (Moreno-López et al., 10/2012). Specifically, we found higher Viso-NODDI values in the supramarginal, prefrontal cortex, posterior cingulate (executive functioning and craving),, lateral occipital and inferior parietal, orbitofrontal (salience attribution and response inhibition), inferior frontal gyrus (cognitive flexibility), dorsolateral, middle frontal (support decision making and inhibition), postcentral (habitation responses), inferior and middle temporal (sensorial recognition), parahippocampal (decision involving memory) (Barrós-Loscertales et al., 2011; Moreno-López et al., 10/2012).

We did not find differences in other regions commonly reported such as amygdala (stress and reward processing), insula (interoception and awareness in SUDs), anterior cingulate and accumbens in striatum (regulation of dopamine via projections to frontal, temporal and limbic cortices) (Barrós-Loscertales et al., 2011). This discrepancy between ordinary volume effects and Viso-NODDI effects could arise from the higher specificity of Viso-NODDI values. As such, the macrostructural volume changes commonly found in these regions may instead reflect adaptive changes and not microstructural pathology (Sierakowiak et al., 2015). However, this would need to be confirmed with animal or human post-mortem studies.

### Clinical relevance of the free-water pathology

Notably, age and years of cocaine use were highly correlated and therefore difficult to disentangle from the analyses. The correlation analysis between Viso-NODDI and clinical measures showed a positive correlation between cocaine age onset with left lateral orbitofrontal and left inferior frontal gyrus pars triangularis, meaning the older they started cocaine use, the worse the pathology. Multiple regression analysis, which considered the contribution of all other demographic and clinical variables in the correlation, showed that left orbitofrontal cortex Viso-NODDI values were only significantly correlated with age and cognitive impulsivity. Moreover, the left inferior frontal gyrus pars triangularis analysis showed it was affected significantly by only cocaine age onset. The positive correlation between cocaine age onset, impulsivity and Viso-NODDI in these regions, may suggest that cocaine use at an early age could be benefited with certain protective mechanisms, which has been found in adolescent rodent models of cocaine use compared to adult models (Kantak, 2020). This needs to be explored further by other studies. Orbitofrontal cortex and inferior frontal gyrus pars triangularis are both integrated in the executive control network involved in response inhibition, working memory and executive function processes (Shen et al., 2020). In general, orbitofrontal cortex pathology seems to be associated with malfunction of motivation value of rewards, impulsive regulation and compulsivity, which, along with the striatum, modulates behavior in the presence of rewards that could be critical for cocaine wanting (Dalley et al., 2011b). The inferior frontal gyrus has been found with volume changes presumably due to cocaine consumption and linked to the continuous consumption (Pando-Naude et al., 2020).

Meanwhile, cocaine dose per week was negatively correlated with left Uncinate, left superior longitudinal fasciculus tracts, and posterior cingulate gyrus, suggesting higher cocaine dosage per week correlated with lower pathology. The multiple regression confirmed this finding in left uncinate and left SLF, while the posterior cingulate was instead significant only with sex and cognitive impulsivity. While the group comparison found higher Viso-NODDI in the cocaine use disorder group, these negative correlations in white matter suggest the pathology may be less related to free-water and perhaps other mechanisms or effects are involved in continuous higher dosage of cocaine such as fiber thinning. Another explanation may be that we do not have enough sample size in the extreme values of cocaine dosage to truly represent the underlying pathology, as the measure of dosage is discrete (see Supplementary Table 8). Nevertheless, without a doubt these regions are affected in cocaine use disorder.

Impulsiveness or impulsivity is a multidimensional construct that is a marker of severity in substance use disorders (Vassileva & Conrod, 2019) and has been described as a behavioral trait and a vulnerability factor that could be part of the personality profile that antecede to starting a drug consumption and contribute to the dependence phenotype (Morein-Zamir & Robbins, 2015). We found that total impulsiveness was positively correlated with right lateral occipital cortex, posterior cingulate gyrus and right postcentral gyrus. The sub-scales of the impulsiveness scale: cognitive and motor impulsivity, where correlated with different regions as confirmed by the multiple regression analysis. Cognitive impulsivity was correlated with posterior cingulate and left orbitofrontal cortex while motor impulsivity was correlated left inferior frontal gyrus pars opercularis, right middle frontal gyrus, right lateral occipital cortex, and right postcentral gyrus. Other studies have found a relationship between impulsiveness and structural pathology (Meade et al., 2020). From the perspective of drug-taking as a product of self-regulatory alteration, finding this association may help to explain why cocaine use persists over the risks involved in the seek for the substance (Argyriou et al., 2018). Postcentral gyrus is a somatosensory region which reveals higher neural activity in motor execution, and when failure to inhibit a motor response occurs, it is associated with an increase of self-reported impulsivity (Herman & Duka, 2019). Correlation with other areas related to substance use suggests impulsiveness as an important trait in cocaine use disorder and structural pathology. Finally, the multiple regression analysis showed that the right occipital lateral cortex was correlated with general drug use while the left inferior frontal gyrus pars opercularis was correlated with psychiatric symptoms (ASIP), which are usually related to substance use disorders (Narvaez et al., 2014).

## Limitations

Viso-NODDI metrics use a bi-tensor model to represent just a single fiber population, however, studies have described that free-water contribution may be not fully estimated since WM voxels have at least two fiber populations in 66 to 90 percent of the time. This may lead to competing signals over these fibers, which are then incorrectly assigned to the free-water compartment. For better fitting, more shells could be necessary to obtain more accurate Viso-NODDI values (Dumont et al., 2019). Variables in the clinical measures such as cocaine dose per week, craving and years of consumption are self-reported, therefore they are subjective measures. However, studies have reported the reliability of these measures for assessing aspects of the pattern of drug use (Secades-Villa & Fernández-Hermida, 2003). The lack of a higher sample of females in the study also limits the generalizability of our results.

## Conclusions

In this study, we found higher Viso-NODDI in cocaine use disorder patients in general, that could suggest microstructural tissue degeneration with extracellular fluids that compound the interconnections of cortical regions between frontal hemispheres and with parietal and temporal lobes which is observed by impairments in impulsivity, motivation, reward and decision-making processes commonly observed in consumers (Grussu et al., 2017; Lim et al., 2008). We also found Viso-NODDI values in these regions correlated with cocaine use severity, onset age of use and impulsivity. More work of microstructure and cellular corroboration are needed to elucidate the relation between free-water changes and neurodegenerative processes such as neuroinflammation in order to develop a better understanding of cocaine use disorder.

## Data Availability

Raw data will be available soon at openneuro. Code and workflow are available in: https://jalilrt.github.io/Viso-NODDI-in-CUD/

https://jalilrt.github.io/Viso-NODDI-in-CUD/

## Acknowledgements

We thank the people who helped this project in one way or another: Vinoo Alluri, Thania Balducci Garcia, Ernesto Reyes Zamorano, Jorge J. Gonzalez Olvera, Francisco J. Pellicer Graham, Margarita López-Titla, Aline Leduc, Erik Morelos-Santana, Diego Angeles Valdez, Alely Valencia, Lya Paas, Daniela Casillas, Sarael Alcauter, Luis Concha and Bernd Foerster. We also thank Rocio Estrada Ordoñez and Isabel Lizarindari Espinosa Luna at the Unidad de Atención Toxicológica Xochimilco for all their help and effort. Finally, we thank the study participants for their cooperation and patience. We also thank the Laboratorio Nacional de Visualización Científica Avanzada (LAVIS) for the use of their computer cluster and the Laboratorio Nacional de Imagenología por Resonancia Magnética (LANIREM). This project was funded by CONACYT-FOSISS project No. 0201493 and CONACYT-Cátedras project No. 2358948.

## Open science

Raw data is available at openneuro (https://openneuro.org/datasets/ds003037/versions/1.0.0). Code and workflow are available in: https://jalilrt.github.io/Viso-NODDI-in-CUD/

## Authors contribution

GVE conceived the experiments; GVE and IM, designed the analysis; SA, RTJ analyzed the data; RTJ and GVE wrote the manuscript; GVE and IM supervised the entire process, and all authors contributed to interpretation of data, revised the manuscript and approved the final manuscript.

## Notes

### Competing Interest Statement

The authors have declared no competing interest.

### Funding Statement

This project was funded by CONACYT-FOSISS project No. 0201493 and CONACYT-Catedras project No. 2358948.

### Author Declarations

The study was approved by the local ethics committee and carried out at the Instituto Nacional de Psiquiatria Ramon de la Fuente Muniz in Mexico City, Mexico, according to the Declaration of Helsinki. All participants were invited through posters placed in several centers for SUD treatment. HCs were recruited from the Institute (i.e. administrative workers) and by fliers around the city. Participants were fully briefed and they provided verbal and written informed consent.

## References

Argyriou, E., Um, M., Carron, C., & Cyders, M. A. (2018). Age and impulsive behavior in drug addiction: A review of past research and future directions. Pharmacology, Biochemistry, and Behavior, 164, 106–117.

Avants, B. B., Epstein, C. L., Grossman, M., & Gee, J. C. (2008). Symmetric diffeomorphic image registration with cross-correlation: evaluating automated labeling of elderly and neurodegenerative brain. Medical Image Analysis, 12(1), 26–41.

Azadeh, S., Hobbs, B. P., Ma, L., Nielsen, D. A., Gerard Moeller, F., & Baladandayuthapani, V. (2016). Integrative Bayesian analysis of neuroimaging-genetic data with application to cocaine dependence. NeuroImage, 125, 813–824.

Barrós-Loscertales, A., Garavan, H., Bustamante, J. C., Ventura-Campos, N., Llopis, J. J., Belloch, V., Parcet, M. A., & Avila, C. (2011). Reduced striatal volume in cocaine-dependent patients. NeuroImage, 56(3), 1021–1026.

Beard, C. L., Schmitz, J. M., Soder, H. E., Suchting, R., Yoon, J. H., Hasan, K. M., Narayana, P. A., Moeller, F. G., & Lane, S. D. (2019). Regional differences in white matter integrity in stimulant use disorders: A meta-analysis of diffusion tensor imaging studies. Drug and Alcohol Dependence, 201, 29–37.

Bell, R. P., Foxe, J. J., Nierenberg, J., Hoptman, M. J., & Garavan, H. (2010). Assessing white matter integrity as a function of abstinence duration in former cocaine-dependent individuals. Drug and Alcohol Dependence. https://doi.org/10.1016/j.drugalcdep.2010.10.001

Burkovetskaya, M. E., Small, R., Guo, L., Buch, S., & Guo, M.-L. (2020). Cocaine self-administration differentially activates microglia in the mouse brain. Neuroscience Letters, 728, 134951.

Dalley, J. W., Everitt, B. J., & Robbins, T. W. (2011). Impulsivity, compulsivity, and top-down cognitive control. Neuron, 69(4), 680–694.

Di Biase, M. A., Katabi, G., Piontkewitz, Y., Cetin-Karayumak, S., Weiner, I., & Pasternak, O. (2020). Increased extracellular free-water in adult male rats following in utero exposure to maternal immune activation. Brain, Behavior, and Immunity, 83, 283–287.

Downey, L. E., Mahoney, C. J., Buckley, A. H., Golden, H. L., Henley, S. M., Schmitz, N., Schott, J. M., Simpson, I. J., Ourselin, S., Fox, N. C., Crutch, S. J., & Warren, J. D. (2015). White matter tract signatures of impaired social cognition in frontotemporal lobar degeneration. NeuroImage: Clinical, 8, 640–651.

Dumont, M., Roy, M., Jodoin, P.-M., Morency, F. C., Houde, J.-C., Xie, Z., Bauer, C., Samad, T. A., Van Dijk, K. R. A., Goodman, J. A., Descoteaux, M., & Alzheimer’s Disease Neuroimaging Initiative. (2019). Free Water in White Matter Differentiates MCI and AD From Control Subjects. Frontiers in Aging Neuroscience, 11, 1.

Fischl, B. (2012). FreeSurfer. NeuroImage, 62(2), 774–781.

Garza-Villarreal, E. A., Chakravarty, M. M., Hansen, B., Eskildsen, S. F., Devenyi, G. A., Castillo-Padilla, D., Balducci, T., Reyes-Zamorano, E., Jespersen, S. N., Perez-Palacios, P., Patel, R., & Gonzalez-Olvera, J. J. (2017). The effect of crack cocaine addiction and age on the microstructure and morphology of the human striatum and thalamus using shape analysis and fast diffusion kurtosis imaging. Translational Psychiatry, 7(5), e1122.

Gorelick, D. A. (2015). Treatment of Cocaine Addiction. In Textbook of Addiction Treatment: International Perspectives (pp. 381–404). https://doi.org/10.1007/978-88-470-5322-9_15

Greicius, M. D., Supekar, K., Menon, V., & Dougherty, R. F. (2009). Resting-state functional connectivity reflects structural connectivity in the default mode network. Cerebral Cortex, 19(1), 72–78.

Grussu, F., Schneider, T., Tur, C., Yates, R. L., Tachrount, M., Ianu$, A., Yiannakas, M. C., Newcombe, J., Zhang, H., Alexander, D. C., DeLuca, G. C., & Gandini Wheeler-Kingshott, C. A. M. (2017). Neurite dispersion: a new marker of multiple sclerosis spinal cord pathology? Annals of Clinical and Translational Neurology, 4(9), 663–679.

Hampton, W. H., Hanik, I. M., & Olson, I. R. (2019). Substance abuse and white matter: Findings, limitations, and future of diffusion tensor imaging research. Drug and Alcohol Dependence, 197, 288–298.

He, Q., Li, D., Turel, O., Bechara, A., & Hser, Y.-I. (2020). White matter integrity alternations associated with cocaine dependence and long-term abstinence: Preliminary findings. Behavioural Brain Research, 379, 112388.

Herman, A. M., & Duka, T. (2019). Facets of impulsivity and alcohol use: What role do emotions play? Neuroscience and Biobehavioral Reviews, 106, 202–216.

Kantak, K. M. (2020). Adolescent-onset vs. adult-onset cocaine use: Impact on cognitive functioning in animal models and opportunities for translation. Pharmacology, Biochemistry, and Behavior, 196, 172994.

Lim, K. O., Wozniak, J. R., Mueller, B. A., Franc, D. T., Specker, S. M., Rodriguez, C. P., Silverman, A. B., & Rotrosen, J. P. (2008). Brain macrostructural and microstructural abnormalities in cocaine dependence. Drug and Alcohol Dependence, 92(1-3), 164–172.

Maillard, P., Fletcher, E., Singh, B., Martinez, O., Johnson, D. K., Olichney, J. M., Farias, S. T., & DeCarli, C. (2019). Cerebral white matter free water: A sensitive biomarker of cognition and function. Neurology, 92(19), e2221–e2231.

Makris, N., Kennedy, D. N., McInerney, S., Sorensen, A. G., Wang, R., Caviness, V. S., Jr, & Pandya, D. N. (2005). Segmentation of Subcomponents within the Superior Longitudinal Fascicle in Humans: A Quantitative, In Vivo, DT-MRI Study. Cerebral Cortex, 15(6), 854–869.

Ma, L., Hasan, K. M., Steinberg, J. L., Narayana, P. A., Lane, S. D., Zuniga, E. A., Kramer, L. A., & Moeller, F. G. (10/2009). Diffusion tensor imaging in cocaine dependence: Regional effects of cocaine on corpus callosum and effect of cocaine administration route. Drug and Alcohol Dependence, 104(3), 262–267.

McLellan, A. T., Luborsky, L., Cacciola, J., Griffith, J., Evans, F., Barr, H. L., & O’Brien, C. P. (1985). New data from the Addiction Severity Index. Reliability and validity in three centers. The Journal of Nervous and Mental Disease, 173(7), 412–423.

Meade, C. S., Bell, R. P., Towe, S. L., & Hall, S. A. (2020). Cocaine-related alterations in fronto-parietal gray matter volume correlate with trait and behavioral impulsivity. Drug and Alcohol Dependence, 206, 107757.

Mitchell, T., Archer, D. B., Chu, W. T., Coombes, S. A., Lai, S., Wilkes, B. J., McFarland, N. R., Okun, M. S., Black, M. L., Herschel, E., Simuni, T., Comella, C., Xie, T., Li, H., Parrish, T. B., Kurani, A. S., Corcos, D. M., & Vaillancourt, D. E. (2019). Neurite orientation dispersion and density imaging (NODDI) and free-water imaging in Parkinsonism. Human Brain Mapping, 40(17), 5094–5107.

Morein-Zamir, S., & Robbins, T. W. (2015). Fronto-striatal circuits in response-inhibition: Relevance to addiction. Brain Research, 1628(Pt A), 117–129.

Moreno-López, L., Catena, A., Fernández-Serrano, M. J., Delgado-Rico, E., Stamatakis, E. A., Pérez-García, M., & Verdejo-García, A. (10/2012). Trait impulsivity and prefrontal gray matter reductions in cocaine dependent individuals. Drug and Alcohol Dependence, 125(3), 208–214.

Morie, K. P., Yip, S. W., Zhai, Z. W., Xu, J., Hamilton, K. R., Sinha, R., Mayes, L. C., & Potenza, M. N. (2017). White-matter crossing-fiber microstructure in adolescents prenatally exposed to cocaine. Drug and Alcohol Dependence, 174, 23–29.

Narayana, P. A., Datta, S., Tao, G., Steinberg, J. L., & Moeller, F. G. (10/2010). Effect of cocaine on structural changes in brain: MRI volumetry using tensor-based morphometry. Drug and Alcohol Dependence, 111(3), 191–199.

Narvaez, J. C. M., Jansen, K., Pinheiro, R. T., Kapczinski, F., Silva, R. A., Pechansky, F., & Magalhães, P. V. (2014). Psychiatric and substance-use comorbidities associated with lifetime crack cocaine use in young adults in the general population. Comprehensive Psychiatry, 55(6), 1369–1376.

Oishi, K., Faria, A. V., Hsu, J., Tippett, D., Mori, S., & Hillis, A. E. (2015). Critical role of the right uncinate fasciculus in emotional empathy. Annals of Neurology, 77(1), 68–74.

Paliwal, P., Hyman, S. M., & Sinha, R. (2008). Craving predicts time to cocaine relapse: further validation of the Now and Brief versions of the cocaine craving questionnaire. Drug and Alcohol Dependence, 93(3), 252–259.

Pando-Naude, V., Toxto, S., Fernandez-Lozano, S., Parsons, E. C., Alcauter, S., & Garza-Villarreal, E. A. (2020). Gray and white matter morphology in substance use disorders: A neuroimaging systematic review and meta-analysis. In Neuroscience (No. biorxiv;2020.05.29.122812v1; p. 2115). bioRxiv.

Reese, E. D., Yi, J. Y., McKay, K. G., Stein, E. A., Ross, T. J., & Daughters, S. B. (2019). Triple Network Resting State Connectivity Predicts Distress Tolerance and Is Associated with Cocaine Use. Journal of Clinical Medicine Research, 8(12). https://doi.org/10.3390/jcm8122135

Reise, S. P., Moore, T. M., Sabb, F. W., Brown, A. K., & London, E. D. (2013). The Barratt Impulsiveness Scale-11: reassessment of its structure in a community sample. Psychological Assessment, 25(2), 631–642.

Romero, M. J., Asensio, S., Palau, C., Sanchez, A., & Romero, F. J. (1/2010). Cocaine addiction: Diffusion tensor imaging study of the inferior frontal and anterior cingulate white matter. Psychiatry Research: Neuroimaging, 181(1), 57–63.

Secades-Villa, R., & Fernández-Hermida, J. R. (2003). The validity of self-reports in a follow-up study with drug addicts. Addictive Behaviors, 28(6), 1175–1182.

Shen, K.-K., Welton, T., Lyon, M., McCorkindale, A. N., Sutherland, G. T., Burnham, S., Fripp, J., Martins, R., & Grieve, S. M. (2020). Structural core of the executive control network: A high angular resolution diffusion MRI study. Human Brain Mapping, 41(5), 1226–1236.

Siciliano, C. A., Ferris, M. J., & Jones, S. R. (08/2015). Cocaine self-administration disrupts mesolimbic dopamine circuit function and attenuates dopaminergic responsiveness to cocaine. The European Journal of Neuroscience, 42(4), 2091–2096.

Sierakowiak, A., Mattsson, A., Gómez-Galán, M., Feminía, T., Graae, L., Aski, S. N., Damberg, P., Lindskog, M., Brené, S., & Åberg, E. (2015). Hippocampal Morphology in a Rat Model of Depression: The Effects of Physical Activity. The Open Neuroimaging Journal, 9(1), 1–6.

Smith, H. R., Beveridge, T. J. R., Nader, S. H., Nader, M. A., & Porrino, L. J. (2019). Regional elevations in microglial activation and cerebral glucose utilization in frontal white matter tracts of rhesus monkeys following prolonged cocaine self-administration. Brain Structure & Function, 224(4), 1417–1428.

van Son, D., Wiers, R. W., Catena, A., Perez-Garcia, M., & Verdejo-García, A. (2016). White matter disruptions in male cocaine polysubstance users: Associations with severity of drug use and duration of abstinence. Drug and Alcohol Dependence, 168, 247–254.

Vassileva, J., & Conrod, P. J. (2019). Impulsivities and addictions: a multidimensional integrative framework informing assessment and interventions for substance use disorders. Philosophical Transactions of the Royal Society of London. Series B, Biological Sciences, 374(1766), 20180137.

Volkow, N. D., Michaelides, M., & Baler, R. (2019). The Neuroscience of Drug Reward and Addiction. Physiological Reviews, 99(4), 2115–2140.

Wilcox, C. E., Teshiba, T. M., Merideth, F., Ling, J., & Mayer, A. R. (2011). Enhanced cue reactivity and fronto-striatal functional connectivity in cocaine use disorders. Drug and Alcohol Dependence, 115(1-2), 137–144.

Xu, J., DeVito, E. E., Worhunsky, P. D., Carroll, K. M., Rounsaville, B. J., & Potenza, M. N. (2010). White matter integrity is associated with treatment outcome measures in cocaine dependence. Neuropsychopharmacology: Official Publication of the American College of Neuropsychopharmacology, 35(7), 1541–1549.

Zhang, H., Schneider, T., Wheeler-Kingshott, C. A., & Alexander, D. C. (2012). NODDI: practical in vivo neurite orientation dispersion and density imaging of the human brain. NeuroImage, 61(4), 1000–1016.

Zhang, J., Gao, J., Shi, H., Huang, B., Wang, X., Situ, W., Cai, W., Yi, J., Zhu, X., & Yao, S. (2014). Sex differences of uncinate fasciculus structural connectivity in individuals with conduct disorder. BioMed Research International, 2014, 673165.

