## Supplementaries figures and tables for "Neurite orientation dispersion and density imaging in cocaine use disorder"

**Support figures:**

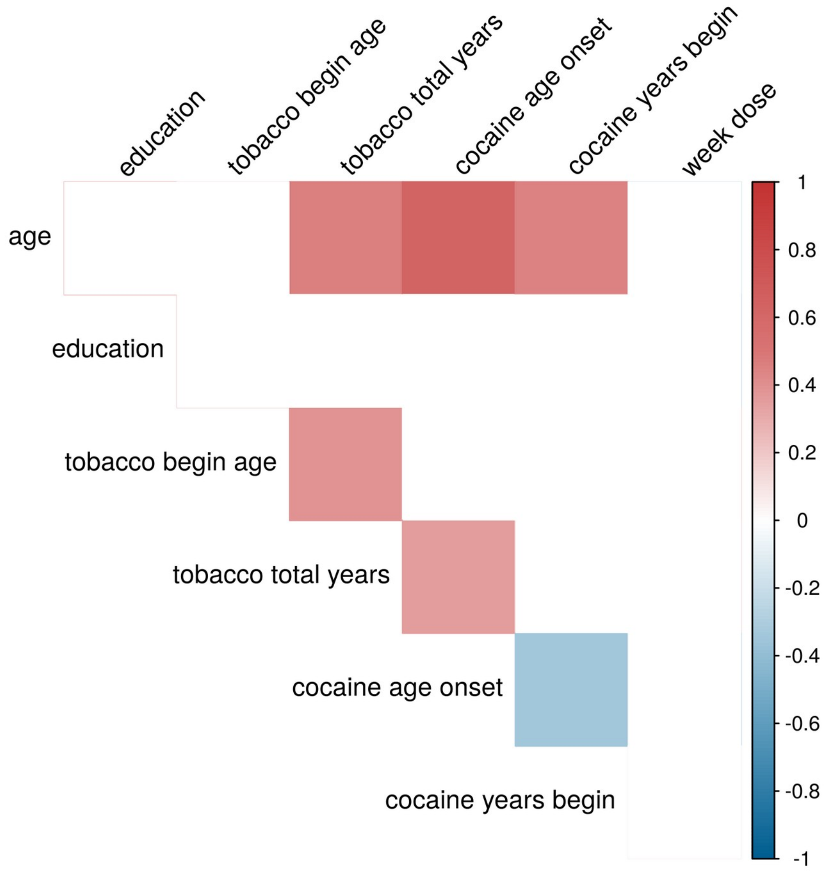


**Figure S1.** Correlation plot among demographic data and drug pattern use, corrected by FDR ($p < 0.05$)

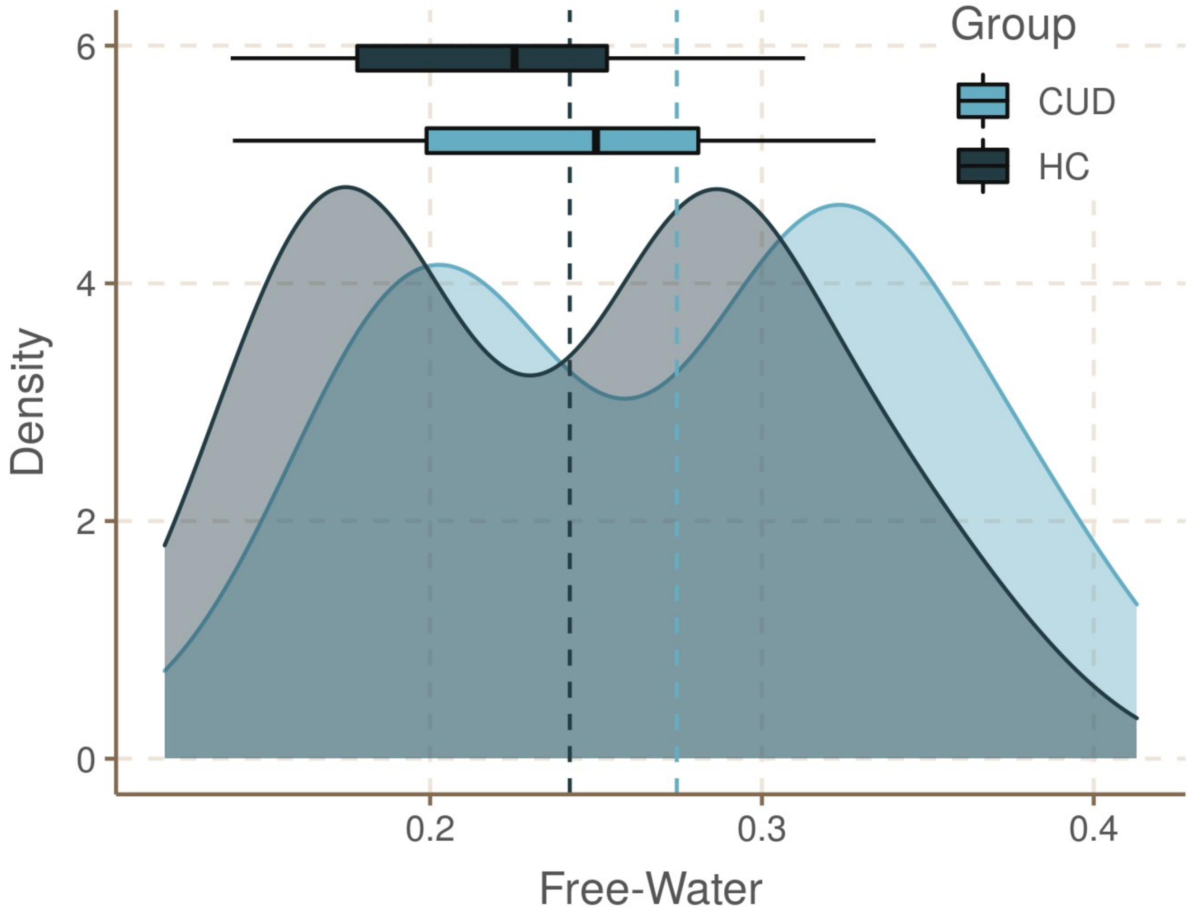


**Figure S2.** Distribution of free-water values in each region of interest (WM-Track and GM) for Cocaine Use Disorder (CUD) and Healthy Controls (HC). Lines represent the mean of each group.

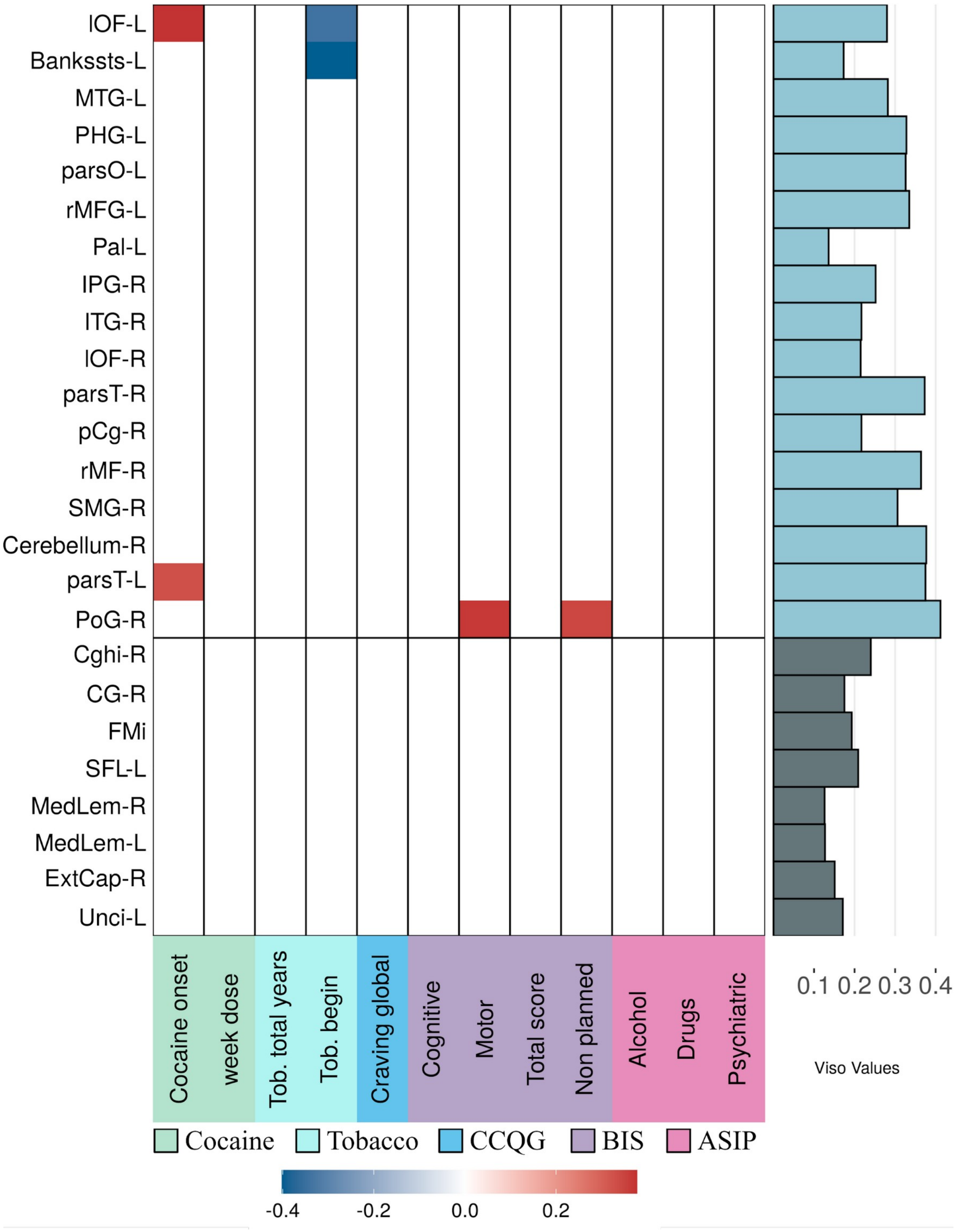


**Figure S3.** Correlation map between significant structures and clinical measures. Blank space means no significance corrected after FDR (p<0.05). Viso-NODDI values of each structure are plotted on the right side. Abbreviation: Cocaine Current Questionnaire (CCQG), Barratt Impulsiveness Scale (BIS), Addiction Severity Index (ASIP).

**Support Tables.**

**Table S1.** Recruitment criteria

| **Inclusion** |
|---|
| Age between 18 – 50 years (CUD/HC) |
| Any sex (CUD/HC) |
| BMI ≧18.5 and ≦ 30.5 (CUD/HC) |
| Cocaine dependency with an active consumption of at least twice a week in the last month (CUD) |
| Desire to participate and agree to the informed consent (CUD/HC) |
| Willingness to cooperate during the MRI sessions (CUD/HC) |

| **Exclusion (all)** |
|---|
| Chronic, cardiovascular and neurological diseases |
| Chronic, cardiovascular and neurological diseases |
| Alcohol dependency |
| MRI contraindications |
| Severe suicide risk |

| **Elimination (all)** |
|---|
| Desire to terminate their participation |
| Abnormal MRI findings |

*CUD = Cocaine Use Disorder, HC = Healthy Control, BMI = Body Mass Index.*

**Table S2.** Clinical instruments

| Instruments | For Criteria |
|---|---|
| MINI-Plus | x |
| Addiction Severity Index (ASI) | |
| Diagnostic Interview for Borderlines-revised, DIB-R | |
| Clinical Global Impression Scale for Borderline Personality Disorder (CGI-BPD) | |
| Edinburgh Handedness Inventory | |
| AMAI NSE 8x7 rule questionnaire | |
| Structured clinical interview for DSM Axis II, self report (SCID-II) | |
| Symptom Checklist-90-revised (SCL-90-R) | |
| Barratt Impulsiveness Scale-11 (BIS-11) | |
| Difficulties in Emotion Regulation Scale (DERS) | |
| Timeline follow back | |
| WHO Disability Assessment Schedule (WHODAS) 2.0 | |
| Cocaine Craving Questionnaire, general and now (CCQ) | |
| Demographic data form, self report | |
| Clinical Interview | x |

*For Criteria = Instruments used for inclusion and exclusion criteria.*

**Table S3.** Psychiatric comorbidities of cocaine addicts.

| Psychiatric Comorbidities | Count (n = 51) |
|---|---|
| Major Depressive Episode current (2 weeks) | 4 |
| Major Depressive Episode Recurrent | 4 |
| Substance Induced Mood Disorder | 7 |
| Major Depressive Episode with Melancholy | 2 |
| Suicidality current (past month) | 13 |
| Suicidality past | 2 |
| Anxiety Disorder with panic due to a general medical condition current | 1 |
| Social Phobia current (past month) | 2 |
| Specific Phobia current | 3 |
| Post-traumatic Stress Disorder current (past month) | 3 |
| Alcoholic dependence (past 12 months) | 13 |
| Alcohol dependence lifetime | 17 |
| Alcohol abuse (past 12 months) | 17 |
| Alcohol abuse lifetime | 22 |
| Substance dependence (non alcohol; past 12 months) | 31 |
| Substance dependence (non alcohol; lifetime) | 32 |
| Generalized anxiety disorder current (past 6 months) | 5 |
| Substance induced generalized anxiety disorder current | 1 |
| Antisocial Personality Disorder lifetime | 10 |
| Somatization Disorder lifetime | 1 |
| Somatization Disorder current | 1 |
| Hypochondriasis current | 1 |
| Conduct Disorder (past 12 months) | 1 |
| Attention Deficit/ Hyperactivity Disorder (past 6 months) | 12 |
| Attention Deficit/ Hyperactivity Disorder (children adolescents; current) | 2 |
| Attention Deficit/ Hyperactivity Disorder (children adolescents; lifetime) | 2 |

| | |
|---|---|
| Attention Deficit/ Hyperactivity Disorder (adults; lifetime) | 13 |
| Attention Deficit/ Hyperactivity Disorder (adults; current) | 13 |
| Adjustment Disorders current | 1 |

*Comorbidities extracted from the Mini-PLUS interview in Spanish version 5.0.0*

**Table S4.** Lifetime medication of cocaine addicts.

| Medication | Count (n = 60) |
|---|---|
| Valproic Acid | 5 |
| Fluoxetine | 7 |
| Pregabaline | 2 |
| Diazepam | 1 |
| Clonazepam | 2 |
| Hydroxyzine | 1 |
| Gabapentine | 1 |
| Topiramate | 3 |
| Risperidone | 1 |
| Sertraline | 1 |
| Olanzapine | 1 |
| Oxcarbazepine | 1 |
| Not Specified | 1 |

Based on patients’ reports in the general interview and the Addiction Severity Index.

**Table S5.** Lifetime history of substance use and abuse.

| Substance | Count (n = 60) |
|---|---|
| Any use of alcohol | 52 |
| Alcohol to intoxication | 40 |
| Heroin | 2 |
| Methadone | 0 |
| Others opiates/analgesic | 2 |
| Barbiturates | 1 |
| Other sedative/hypnotics/tranquilizers | 2 |
| Cocaine | 59 |
| Amphetamines | 5 |
| Cannabis | 21 |
| Hallucinogens | 7 |
| Inhalants | 2 |

The table was created from the Addiction Severity Index items D1-D9.

**Table S6.** Comparison of isotropic volume fraction of extracellular fluid (Viso-NODDI) between Cocaine Use Disorder (CUD) and Healthy Controls (HC) in White Matter (WM) tracts.

| WM Tracts | CUD | HC | p-Value |
|---|---|---|---|
| Cingulum (cingulate gyrus) R (Cg-R) | 0.174 ± 0.057 | 0.149 ± 0.035 | 0.021* |
| Cingulum (hippocampus gyrus) R (Cghi-R) | 0.207 ± 0.052 | 0.241 ± 0.064 | 0.01* |
| Forceps minor (FMi) | 0.193 ± 0.035 | 0.177 ± 0.022 | 0.012* |
| Superior longitudinal fasciculus L (SLF-L) | 0.209 ± 0.036 | 0.19 ± 0.029 | 0.012* |
| Corticospinal tract L (CST-L) | 0.178 ± 0.035 | 0.168 ± 0.026 | 0.148 |
| Corticospinal tract R (CST-R) | 0.172 ± 0.037 | 0.159 ± 0.024 | 0.072 |
| Inferior fronto-occipital fasciculus R (ILOF-R) | 0.165 ± 0.036 | 0.154 ± 0.022 | 0.105 |
| Inferior longitudinal fasciculus R (ILF-R) | 0.163 ± 0.038 | 0.154 ± 0.021 | 0.227 |
| Superior longitudinal fasciculus R (SLF-R) | 0.179 ± 0.042 | 0.167 ± 0.03 | 0.152 |
| Uncinate fasciculus L (Unci-L) | 0.198 ± 0.044 | 0.188 ± 0.03 | 0.033* |

**Table S7.** Comparison of isotropic volume fraction of extracellular fluid (Viso) between Cocaine Use Disorder (CUD) and Healthy Controls (HC) in Gray Matter (GM) Rois.

| GM ROIs | CUD | HC | p-Value |
|---|---|---|---|
| ctx-rh-posteriorcingulate | 0.216 ± 0.085 | 0.176 ± 0.054 | 0.011* |
| ctx-rh-bankssts | 0.15 ± 0.086 | 0.126 ± 0.049 | 0.122 |
| ctx-rh-caudalanteriorcingulate | 0.221 ± 0.094 | 0.185 ± 0.098 | 0.085 |
| ctx-rh-cuneus | 0.307 ± 0.075 | 0.28 ± 0.073 | 0.096 |
| ctx-rh-entorhinal | 0.334 ± 0.09 | 0.302 ± 0.096 | 0.138 |
| ctx-rh-fusiform | 0.191 ± 0.075 | 0.167 ± 0.052 | 0.063 |
| ctx-rh-inferiorparietal | 0.252 ± 0.074 | 0.226 ± 0.044 | 0.042* |
| ctx-rh-inferiortemporal | 0.217 ± 0.06 | 0.194 ± 0.046 | 0.03* |
| ctx-rh-lateraloccipital | 0.216 ± 0.07 | 0.19 ± 0.046 | 0.031* |
| ctx-rh-middletemporal | 0.277 ± 0.067 | 0.253 ± 0.052 | 0.06 |
| ctx-rh-parahippocampal | 0.3 ± 0.071 | 0.271 ± 0.087 | 0.08 |
| ctx-rh-parsopercularis | 0.317 ± 0.096 | 0.291 ± 0.069 | 0.186 |
| ctx-rh-parsorbitalis | 0.312 ± 0.086 | 0.281 ± 0.078 | 0.085 |
| Ctx-rh-parstriangularis | 0.374 ± 0.086 | 0.337 ± 0.071 | 0.038* |
| ctx-rh-pericalcarine | 0.301 ± 0.071 | 0.274 ± 0.065 | 0.08 |
| ctx-rh-precentral | 0.38 ± 0.074 | 0.35 ± 0.072 | 0.052 |
| ctx-rh-rostralanteriorcingulate | 0.235 ± 0.07 | 0.21 ± 0.044 | 0.063 |
| ctx-rh-rostralmiddlefrontal | 0.363 ± 0.079 | 0.321 ± 0.083 | 0.018* |
| ctx-rh-superiorfrontal | 0.353 ± 0.071 | 0.33 ± 0.064 | 0.135 |
| ctx-rh-supramarginal | 0.306 ± 0.073 | 0.272 ± 0.062 | 0.016* |
| ctx-rh-frontalpole | 0.279 ± 0.102 | 0.239 ± 0.092 | 0.065 |
| ctx-rh-transversetemporal | 0.329 ± 0.098 | 0.298 ± 0.106 | 0.166 |
| ctx-rh-insula | 0.32 ± 0.072 | 0.294 ± 0.071 | 0.096 |
| Right-Pallidum | 0.121 ± 0.029 | 0.134 ± 0.039 | 0.093 |
| ctx-lh-bankssts | 0.172 ± 0.084 | 0.141 ± 0.054 | 0.042* |

| | | | |
|---|---|---|---|
| ctx-lh-entorhinal | 0.337 ± 0.071 | 0.032 ± 0.104 | 0.632 |
| ctx-lh-fusiform | 0.201 ± 0.073 | 0.184 ± 0.06 | 0.174 |
| ctx-lh-inferiortemporal | 0.213 ± 0.066 | 0.198 ± 0.045 | 0.22 |
| ctx-lh-lateraloccipital | 0.228 ± 0.074 | 0.205 ± 0.054 | 0.103 |
| ctx-lh-lateralorbitofrontal | 0.28 ± 0.061 | 0.253 ± 0.046 | 0.016* |
| ctx-lh-middletemporal | 0.281 ± 0.071 | 0.248 ± 0.056 | 0.018* |
| Ctx-lh-parahippocampal | 0.329 ± 0.068 | 0.297 ± 0.079 | 0.043* |
| ctx-lh-parsopercularis | 0.325 ± 0.076 | 0.294 ± 0.065 | 0.042* |
| ctx-lh-parsorbitalis | 0.343 ± 0.093 | 0.314 ± 0.076 | 0.099 |
| ctx-lh-parstriangularis | 0.374 ± 0.076 | 0.334 ± 0.07 | 0.012* |
| ctx-lh-postcentral | 0.408 ± 0.068 | 0.38 ± 0.069 | 0.068 |
| ctx-lh-posteriorcingulate | 0.216 ± 0.08 | 0.186 ± 0.065 | 0.08 |
| ctx-lh-precentral | 0.379 ± 0.069 | 0.353 ± 0.072 | 0.082 |
| ctx-lh-rostralmiddlefrontal | 0.334 ± 0.077 | 0.291 ± 0.084 | 0.014* |
| ctx-lh-superiorfrontal | 0.337 ± 0.074 | 0.312 ± 0.06 | 0.079 |
| ctx-lh-superiortemporal | 0.304 ± 0.07 | 0.282 ± 0.064 | 0.151 |
| ctx-lh-frontalpole | 0.264 ± 0.114 | 0.229 ± 0.086 | 0.095 |
| ctx-lh-insula | 0.317 ± 0.07 | 0.289 ± 0.065 | 0.054 |
| Left-Hippocampus | 0.373 ± 0.051 | 0.354 ± 0.053 | 0.124 |
| Left-Amygdala | 0.183 ± 0.068 | 0.166 ± 0.059 | 0.241 |

**Table S8.** Partial correlations

| Clinical | Region | r | p | pFDR |
|---|---|---|---|---|
| BIS11-Cognitive | postcentral-R | 0.286 | 0.029 | 0.040 |
| | Posterior cingulate-R | 0.312 | 0.017 | 0.040 |
| BIS11-Motor | postcentral-R | 0.379 | 0.003 | 0.029 |
| | superior temporal sulcus-L | 0.274 | 0.037 | 0.041 |
| | IFG pars triangularis-L | 0.265 | 0.043 | 0.046 |
| | IFG pars opercularis-L | 0.283 | 0.03 | 0.040 |
| | rostral middle frontal-L | 0.292 | 0.025 | 0.040 |
| | rostral middle frontal-R | 0.262 | 0.046 | 0.046 |
| | posterior Cingulate-R | 0.303 | 0.04 | 0.020 |
| BIS11-Non planned | postcentral-R | 0.28 | 0.032 | 0.040 |
| BIS11-Total score | Lateral Occipital-R | 0.279 | 0.033 | 0.040 |
| | Posterior Cingulate-R | 0.309 | 0.017 | 0.040 |
| | postcentral-R | 0.353 | 0.006 | 0.039 |
| Cocaine Age onset | Lateral Orbitofrontal-L | 0.405 | 0.001 | 0.028 |
| | IFG pars triangularis-L | 0.333 | 0.010 | 0.040 |
| Cocaine dose per week | Posterior Cingulate-R | -0.294 | 0.024 | 0.040 |
| | Superior Longitudinal Fasciculus-L | -0.283 | 0.030 | 0.040 |
| | Uncinate Fasciculus-L | -0.294 | 0.025 | 0.040 |

Partial correlation among clinical measures with significant structures for the CUD group, controlling for Education and Sex. Abbreviations: r = Pearson's coefficients, p = p-Value, pFDR = p-value after correction.
